# Incidence and risk factors of rectovaginal fistula after rectal cancer surgery:a systematic review and meta-analysis

**DOI:** 10.1101/2025.04.09.25325534

**Authors:** Baoliang Hu, Yifei Zheng, Aohua Ti, Yanan Zhu, Xiaojun Li

## Abstract

**Background:** Rectovaginal fistula (RVF) is a unique complication for women after rectal cancer surgery (RCS). Understanding its incidence and risk factors can help further recognize and prevent the disease. Although there have been studies on the risk factors for RVF after RCS, the results show some heterogeneity and even contradictory conclusions due to differences in trial design, sample size, study population, and treatment methods, etc. However, no relevant meta-analyses have been found to date. This study aims to investigate the incidence and risk factors of RVF after RCS through a meta-analysis.

**Methods:** A systematic literature was conducted in PubMed, Embase, Web of Science, and Cochrane Library from the establishment of the database to June 2024. The quality of the literature was evaluated using the Newcastle-Ottawa scale. Meta-analysis was performed using R Studio 4.3.2 software.

**Results:** A total of 1 randomized controlled study and 7 non-randomized controlled studies were included, involving 4920 patients, among which 134 cases had RVF. The meta-analysis results showed that the incidence of RVF after RCS was 3.2% (95%CI: 1.9% ∼ 7.8%). Neoadjuvant chemoradiotherapy,tumors in lower position,longer surgery duration and double-stapled technique were risk factors for the occurrence of RVF after RCS. Laparoscopic technique was a protective factor for RVF after RCS. T4 stage,age,diabetes,ASA anesthesia classification, Prior hysterectomy, Combined pelvic organ resection, and vaginectomy had no significant effect on RVF.

**Conclusion:** The incidence of RVF after RCS is higher than the general cognition, which should be paid attention to by surgeons. The results of this study may help surgeons identify high-risk inpatients in time, take corresponding measures in advance, and expect to reduce the incidence of the disease.

## 1 Introduction

Rectovaginal fistula (RVF) is an abnormal channel between the rectal and vaginal epithelium. The main clinical manifestations are vaginal exhalation and defecation, accompanied by severely uncontrollable defection disorder^[1]^. Obstetric trauma is the most common cause of RVF, and other causes include gynecological tumors, inflammatory bowel disease, and sexual intercourse^[2–5]^. However, with the widespread adoption of radical surgery for rectal cancer, RVF after rectal cancer surgery (RCS) has become a complication that cannot be ignored^[6]^.It is difficult to treat and prone to recurrence, causing not only physiological pains for patients but also severely affecting their psychological states and social functioning^[7, 8]^. The occurrence of RVF after rectal malignancy is related to various factors, including surgical technique, patient’s basic health status, tumor biological characteristics, and postoperative treatment, etc.^[9]^. Although there are studies on the incidence of RVF and related risk factors after RCS, the results show some heterogeneity and even contradictory conclusions due to differences in trial design, sample size, study population, and treatment methods, etc. Currently, there is no meta-analysis on the incidence and risk factors of RVF after RCS. Therefore, to more accurately assess the incidence of RVF after surgery for rectal malignancies and to identify and summarize potential risk factors, a systematic literature review and meta-analysis are necessary. The aim of this study is to synthesize the results of current published studies through meta-analysis to provide an overall estimate of the incidence of RVF after RCS and to explore possible risk factors. Through this study, we hope to provide clinicians with stronger evidence to guide surgical decision-making, improve preoperative assessment and postoperative management, and ultimately reduce the incidence of RVF and improve patient outcomes.

## 2 Materials and Methods

This Meta-analysis was conducted strictly in accordance with the PRISMA reporting guidelines^[10]^. The protocol has been registered and approved on the PROSPERO website (registration number CRD42024546979). Since this Meta-analysis is based on previously published studies, ethical approval from a review board and informed consent from patients are not required.

### 2.1 Literature Search

PubMed, Embase, Web of Science, and the Cochrane Library were searched from the inception of each database to June 2024. Original literature were obtained by combining subject terms with free-text terms and using Boolean logic operators. The search subject terms were: “rectal neoplasms” and “rectovaginal fistula”; free-text terms: “rectal cancer / carcinoma / tumors” and “rectovaginal fistula / fistulas, rectovaginal / Rectovaginal fistulas”. Additionally, the references of relevant studies and related journals were searched manually to ensure the comprehensiveness of the meta-analysis.

### 2.2 Inclusion and Exclusion Criteria

Inclusion Criteria: (1) Study subjects are patients with pathologically confirmed rectal cancer; (2) Patients undergo RCS; (3) Studies report the occurrence of postoperative RVF; (4) Study types include prospective or retrospective case-control studies, cross-sectional studies, cohort studies, or randomized controlled trials. Exclusion Criteria: (1) Study subjects include other basic conditions unrelated to rectal malignancies, such as anal cancer, gynecological tumors, inflammatory bowel disease, etc.; (2) Studies published in languages other than English, conference abstracts, reviews, commentaries, duplicate publications, and literature with the same data from the same author; (3) Reports with small sample sizes (n < 5); (4) Incomplete data, incomplete descriptions, or data that cannot be extracted or analyzed.

### 2.3 Literature Extraction and Quality Assessment

EndNote 20.6 was used to automatically deduplicate the retrieved original literature. Two researchers independently conducted literature screening and quality assessment, and any discrepancies were resolved through discussion with a third author. After reading the full text, two researchers independently extracted information and then cross-checked it. If information was incomplete, supplementary data was sought through email contact with the authors. The quality of studies was assessed using the “Newcastle-Ottawa Scale (NOS)”^[11]^, which includes the selection of study population, comparability, and exposure or outcome assessment. This scale uses a semi-quantitative star system, with a maximum of 9 stars.NOS score>6 points were considered high quality.

### 2.4 Statistical Methods

Statistical analysis was performed using R Studio 4.3.2 software. The heterogeneity of the results across studies were assessed using *I^2^*. When *I^2^* is *≤* 50%, it indicates good homogeneity among the studies, and a fixed-effect model was applied to combine and analyze the data. Otherwise, a random-effects model was used, and the reasons for heterogeneity were analyzed. Sensitivity analysis was also conducted to test the stability of the results. For dichotomous variables, the odds ratio (OR) was used as the summary statistic, and for continuous data, the mean difference (MD) was used as the summary statistic, with the corresponding 95% confidence intervals (CI) calculated. When the number of included studies was greater than 10, funnel plots were used to analyze publication bias. A P-value of less than 0.05 was considered statistically significant.

## 3 Results

### 3.1 Literature Search and Quality Assessment

After searching various databases, a total of 897 articles were obtained. 563 articles remained after excluding duplicate studies. 478 articles were excluded after reviewing the titles and abstracts. 77 articles were excluded after reading the full texts. Ultimately, 8 articles were included (Fig 1), comprising 6 case-control studies, 1 one arm study, and 1 retrospective cohort, with a total of 4920 patients included. The basic characteristics of the included literature are shown in Table 1.

**Table 1.**
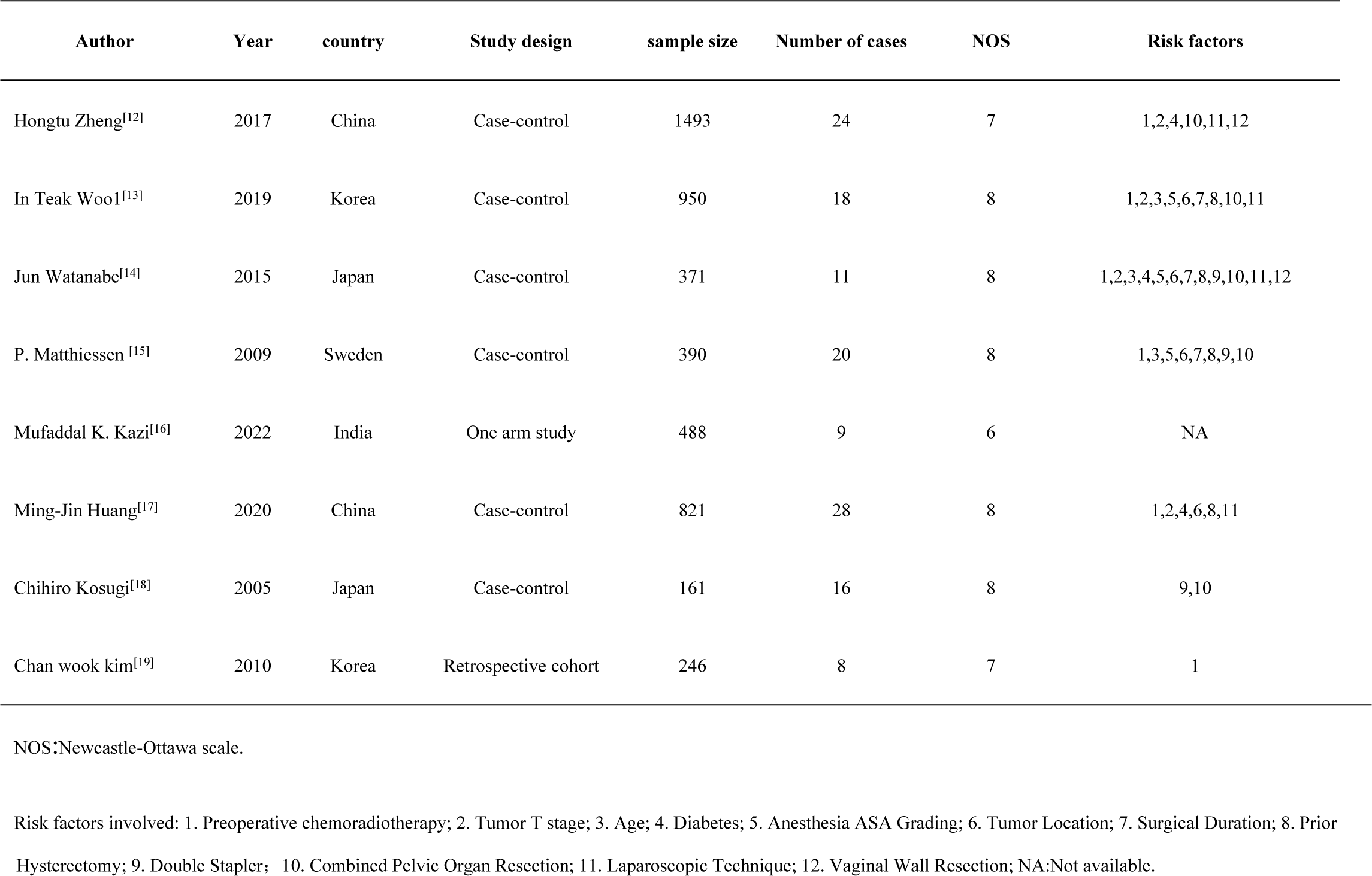
Basic characteristics of included literature.

### 3.2 Meta-analysis on Incidence of RVF

#### 3.2.1 Incidence

A total of 8 articles were included, involving 4920 female rectal cancer patients who underwent surgical treatment, among which 134 cases were RVFs. The results of random-effects model meta-analysis are shown in Fig 2. The overall incidence of RVF after RCS was 3.2% (95%CI: 1.9% ∼ 4.8%).

#### 3.2.2 Sensitivity Analysis

Sensitivity analysis was conducted using the leave-one-out method. The results showed no significant changes in the incidence of RVF, indicating that the findings are stable and reliable (Fig 3).

### 3.3 Risk Factors

#### 3.3.1 Preoperative Chemoradiotherapy

A total of 6 articles examined the impact of preoperative chemoradiotherapy on the occurrence of RVF after RCS. There was significant heterogeneity among these studies (*P* < 0.01, *I^2^* = 80%), prompting a random-effects model was used for the meta-analysis. There was a statistically significant difference between patients who received preoperative chemoradiotherapy and those who did not (OR = 3.72, 95%CI: 1.34∼10.36)(Fig 4).

#### 3.3.2 Tumor T Stage

A total of 4 articles compared the impact of T4 stage versus non-T4 stage on the occurrence of RVF after RCS. There was heterogeneity among the studies (*P* = 0.08, *I^2^* = 56%), hence a random-effects model was used for the meta-analysis. The results showed that there was no statistically significant difference between T4 stage and non-T4 stage (OR = 2.19, 95%CI: 0.99∼ 4.87)(Fig 5).

#### 3.3.3 Age

A total of 5 articles compared the impact of age on RVF. All included studies showed no significant statistical difference in the incidence of RVF with respect to age (*P* > 0.05). Among them, 1 article did not provide detailed data, and 1 article compared the difference in RVF between those over 65 years old and those under 65 years old. Therefore, we analyzed for the remaining 3 articles on the difference in RVF between patients over 75 years old and those under 75 years old. There was no heterogeneity among the 3 studies (P = 0.60, *I^2^* = 0%), so a fixed-effect model was used for the meta-analysis. The results showed that there was no statistically significant difference in RVF between patients over 75 years old and those under 75 years old (OR = 1.00, 95%CI: 0.50-2.00)(Fig 6). This finding suggests that age may not be a significant risk factor for the development of RVF after RCS.

#### 3.3.4 Diabetes

A total of 3 studies compared the impact of diabetes on the occurrence of RVF after RCS. There was no heterogeneity among the studies (*P* = 0.54, *I^2^* = 0%), so a fixed-effect model was used for the meta-analysis. The results showed that there was no statistically significant difference in RVF between patients with diabetes and those without diabetes (OR = 0.42, 95%CI: 0.13-1.33)(Fig 7). This suggests that diabetes may not be a significant risk factor for the development of RVF following RCS.

#### 3.3.5 Anesthesia ASA Grading

A total of 3 studies compared the impact of ASA grades on RVF, dividing patients into those with an ASA grade less than III and those with an ASA grade of III or higher. There was no heterogeneity among the studies (*P* = 0.80, *I^2^* = 0%), so a fixed-effect model was used for the meta-analysis. The results showed that there was no statistically significant difference between patients with an ASA grade of III or higher and those with an ASA grade less than III (OR = 1.34, 95%CI: 0.44 ∼ 4.10)(Fig 8). This suggests that ASA grading may not be a significant risk factor for the development of RVF following RCS.

#### 3.3.6 Tumor Location

A total of 4 studies compared the impact of tumor location on RVF. So we analyzed the effects on RVF between distances from tumor to the anus more than 5 cm and less than 5 cm. There was no heterogeneity among the studies (*P* = 0.79, *I^2^* = 0%), so a fixed-effect model was used for the meta-analysis. The results showed that there was a statistically significant difference in the impact on RVF between tumors less than 5 cm from the anus and those greater than 5 cm (OR = 7.33, 95%CI: 4.16-12.91)(Fig 9). This suggests that tumor location is a significant risk factor for the development of RVF following RCS.

#### 3.3.7 Surgical Duration

A total of 3 studies compared the impact of surgical duration on RVF. Therefore, we analyzed the effects of surgical duration exceeding 240 min versus less than 240 min on RVF. There was no heterogeneity among the studies (*P* = 0.59, *I^2^* = 0%), so a fixed-effect model was used for the meta-analysis. The results showed that there was a statistically significant difference between surgeries duration more than 240 minutes and those less than 240 minutes (OR = 2.35, 95%CI: 1.31-4.20)(Fig 10). This suggests that longer surgical duration may be a risk factor for the development of RVF following RCS.

#### 3.3.8 Prior Hysterectomy

A total of 4 studies compared the impact of prior hysterectomy on RVF. There was no heterogeneity among the studies (*P* = 0.71, *I^2^* = 0%), so a fixed-effect model was used for the meta-analysis. The results showed that there was no statistically significant difference between patients who had undergone hysterectomy and those who had not (OR = 1.78, 95%CI: 0.92-3.46)(Fig 11). This suggests that prior hysterectomy may not be a significant risk factor for the development of RVF following RCS.

#### 3.3.9 Double Stapler

Three studies compared the impact of single versus double stapler techniques on RVF after RCS. There was no heterogeneity among the studies (*P* = 0.66, *I^2^* = 0%), so a fixed-effect model was used for the meta-analysis. The results showed that there was a statistically significant difference in the treatment effect on RVF between the double stapler and single stapler techniques (OR = 2.94, 95%CI: 1.18-7.35)(Fig 12).This suggests that the double stapler technique may be associated with a higher risk of RVF compared to the single stapler technique*。*

#### 3.3.10 Combined pelvic Organ Resection

A total of 2 studies compared the impact of combined pelvic organ resection on RVF. There was significant heterogeneity among the studies (*P* = 0.07, *I^2^* = 69%), hence a random-effects model was used for the meta-analysis. The results showed that the impact of combined pelvic organ resection on RVF was not statistically significant (OR = 4.49, 95%CI: 0.84 ∼ 23.87)(Fig 13).

#### 3.3.11 Laparoscopic Technique

A total of 4 studies compared the impact of laparoscopic surgery versus open surgery on RVF after RCS. There was little heterogeneity among the studies (*P* = 0.24, *I^2^* = 28%), so a fixed-effect model was used for the meta-analysis. The results showed that there was a statistically significant difference in RVF between laparoscopic surgery and open surgery (OR = 0.51, 95%CI: 0.29-0.88)(Fig 14). This suggests that laparoscopic surgery may be associated with a lower risk of RVF compared to open surgery.

#### 3.3.12 Vaginal Wall Resection

Two studies compared the impact of vaginal wall resection on RVF. There was significant heterogeneity among the studies (*P* = 0.05, *I^2^* = 74%), leading to the use of a random-effects model for the meta-analysis. The results showed that there was no statistically significant difference between patients who underwent vaginal wall resection and those who did not (OR = 7.94, 95%CI: 0.64-99.08)(Fig 15).

### 3.4 Publication bias

We generated funnel plots for each risk factor and conducted a publication bias assessment.Visual examination of most of the funnel plots showed asymmetrical distribution, suggesting possible publication bias(Fig 16).This may be due to the small amount of literature.

## 4 Discussion

RVF is one of the serious complications following RCS, characterized by difficult treatment, high costs, suboptimal outcomes, and high recurrence rates. It prolongs patient treatment duration and can have a profound psychological impact on patients, warranting clinical attention and prevention^[9]^. Through meta-analysis, this study identified risk factors for RVF after RCS, including preoperative chemoradiotherapy, prophylactic ostomy, double stapler, surgical duration, tumor location, and laparoscopic technique.

In recent years, with the increasing volume of RCS, the number of patients with postoperative RVF has also risen. However, the reported incidence of RVF after RCS varies across literature^[20]^. The results of this study indicate an incidence rate of 3.2% for RVF after RCS, which aligns with some recent literature^[1]^. Based on the incidence rate found in this study, RVF is not an uncommon complication and should be a concern in clinical practice.

The identification of these risk factors is crucial for clinicians to better understand the potential predictors of RVF and to implement preventive measures. Preoperative chemoradiotherapy, for instance, has been shown to increase the risk, which may guide surgeons to consider alternative treatment strategies or enhanced postoperative care for patients undergoing this therapy. Similarly, the impact of tumor stage, surgical techniques, and other identified factors can inform surgical planning and decision-making to minimize the risk of fistula development.

### 4.1 Preoperative Chemoradiotherapy

Preoperative chemoradiotherapy has been widely accepted as beneficial for the long-term prognosis of rectal cancer and is included in treatment guidelines^[21–23]^. However, neoadjuvant therapy may increase the risk of incision complication^[24]^.Currently, there is a lack of large-sample, high-quality randomized controlled studies on the impact of preoperative chemoradiotherapy on RVF after RCS. Existing studies have yielded inconsistent results. The results of this meta-analysis show that preoperative chemoradiotherapy increases the incidence of postoperative RVF. This may be related to several factors: preoperative chemoradiotherapy can lead to increased local tissue edema, scar hyperplasia, unclear boundaries between the rectum and vagina, and increased surgical difficulty, thereby increasing the risk of RVF^[25, 26]^. Additionally, the impact of preoperative chemoradiotherapy on surgical complications is a concern for surgeons.This suggests that preoperative chemoradiotherapy may have some impact on surgical complications, and the balance between its therapeutic effects and the risk of complications needs to be considered comprehensively in clinical practice.

### 4.3 Tumor Location

The literature has consistently reported that the lower the tumor location, the higher the risk of anastomotic leaks and RVF^[27]^. This is in line with the findings of our study. Generally, the lower the tumor location, the smaller the surgical operating space and the greater the surgical difficulty, which correspondingly increases the probability of vaginal damage. As mentioned in the studies, the lower the tumor is located, the larger the rectal anterior space separation during surgery, which increases the possibility of damaging the vaginal posterior wall^[12]^. Additionally, lower tumor locations result in poorer exposure, increasing the likelihood of including the vaginal posterior wall when using a stapler for anastomosis^[12]^. Furthermore, tumors located less than 5 cm from the anal verge are identified as an independent risk factor for anastomotic leaks, with an increased risk of up to 8 times^[17]^. The reasons for this increased risk may include larger resection surfaces leading to more seroma and bleeding, which can result in poor anastomotic blood supply; the loss of serosal protection after bowel resection, leading to increased infection rates; and the increased tension on the anastomosis due to the removal of a segment of the bowel^[17]^.

### 4.4 Surgical Duration

The results of this study indicate that a surgical duration exceeding 240 minutes is a risk factor for the development of RVF after RCS. This is consistent with the understanding that prolonged surgical duration increases the risk of anastomotic leaks postoperatively^[28]^. Extended surgical duration can lead to increased blood loss and infection risk, and it can also exacerbate tissue edema and ischemia^[29–31]^, which affects local healing and is another reason for the increased risk of RVF. According to a retrospective study analyzing 1123 cases of female RCS patients from 1997 to 2008, the occurrence of RVF was closely related to the duration of surgery, with longer surgery duration being associated with a higher risk of fistula development^[14]^. This highlights the importance of efficient surgical techniques and procedures to minimize the duration of surgery and potentially reduce the risk of postoperative complications such as RVFs.

### 4.5 Double Stapler Technique

A significant amount of literature has reported that the use of double stapler techniques is associated with a higher risk of RVF compared to single stapler techniques^[5, 7, 32]^[5, 33, 34], which aligns with the findings of this study. For low-lying rectal cancers, the surgical space is often limited, making it challenging to fully mobilize the rectum and vagina during surgery. This difficulty can lead to the inclusion of the vagina in the circular stapler, which is not fully mobilized, resulting in RVF^[32, 33]^. The use of double stapler techniques, particularly in the context of lower RCS, has been identified as a risk factor for the development of RVF. This is due to the increased complexity of the surgery and the potential for inclusion of the vaginal wall in the stapler line, which can lead to RVF^[32]^. It is crucial for surgeons to be aware of this risk and to take appropriate measures to minimize the possibility of including the vaginal wall during anastomosis^[33]^.

### 4.6 Laparoscopic Technique

Studies have indicated that laparoscopic techniques can reduce surgical blood loss, minimize trauma, and decrease the incidence of postoperative complications in RCS^[34, 35]^, which is consistent with the findings of this study. Compared to open surgery, laparoscopic techniques can reduce the risk of RVF after RCS. Laparoscopic procedures offer better exposure and clearer vision compared to open surgery, which reduces the risk of accidental vaginal damage. The use of laparoscopic techniques allows for more precise dissection and identification of anatomical planes, which can be particularly beneficial in surgeries involving the rectum and adjacent structures such as the vagina. This precision can help to avoid complications like RVFs. The improved visualization provided by laparoscopy can facilitate better tissue handling and decrease the risk of injury to surrounding organs, including the vagina, thus potentially lowering the risk of postoperative RVF^[34]^.

### 4.7 Limitations of this Study

Firstly, this study only searched for published English-language literature, which may introduce a certain degree of selection bias. Secondly, when extracting potential risk factors that could affect RVF after RCS, we found that some studies did not exclude other confounding factors from potential risk factors, which could affect our results. Thirdly, there is a scarcity of original literature on risk factors for RVF after RCS, especially randomized controlled trials, which can impact the reliability of the outcomes. Lastly, there may be other potential risk factors that were not reported in the included literature.

It is important to acknowledge these limitations when interpreting the results of this meta-analysis. Efforts should be made in future studies to address these limitations, such as expanding the search to include non-English literature, controlling for confounding factors more rigorously, increasing the number of randomized controlled trials, and thoroughly reporting all potential risk factors to provide a more comprehensive understanding of the risks associated with RVF after RCS.

## 5 Outlook

Based on the risk factors for RVF after RCS identified in this study, future research can focus on developing predictive models to assess the risk of RVF following RCS. Given the seriousness of this postoperative complication, it is hoped that more researchers will conduct in-depth studies to provide valuable support for clinical decision-making. The development of predictive models could involve integrating various risk factors, such as preoperative chemoradiotherapy, tumor stages, surgical techniques, and patient-specific factors, to create a comprehensive assessment tool. This tool could help surgeons to identify patients at higher risk for RVF preoperatively and tailor their surgical approaches accordingly.

Additionally, future studies could explore the impact of novel surgical techniques, advancements in anastomotic devices, and postoperative management strategies on the incidence of RVF. Research into the biological and molecular mechanisms underlying the development of fistulas may also lead to new therapeutic targets for prevention and treatment.

Furthermore, multicenter, prospective studies with larger sample sizes and longer follow-up periods are needed to validate the findings of this meta-analysis and to establish evidence-based guidelines for the prevention and management of RVF in rectal cancer patients.

In conclusion, the outlook for future research in this area is to enhance our understanding of the pathogenesis of RVF, improve risk stratification, and optimize clinical practices to reduce the incidence of this devastating complication and improve patient outcomes.

## Data Availability

My paper is a meta-analysis, all the data are from published literature, and all the data are public.

## Abbreviations

RCS: rectal cancer surgery;
RVF: Rectovaginal fistula.

## Ethics approval and consent to participate

Not applicable.

## Funding

The work was supported by Talent Support Program of Shaanxi Provincial People’s Hospital (2023LJ-01).

## Supporting information

PRISMA_2020_checklist.(DOCX)

## Declaration of competing interest

The authors declare no conflicts of interest for this article.

## Author Contributions

Methodology: Baoliang Hu, Yifei Zheng.

Resources: Baoliang Hu, Aohua Ti

Software: Yanan Zhu.

Writing - original draft; Writing -review & editing:Xiaojun Li.

---

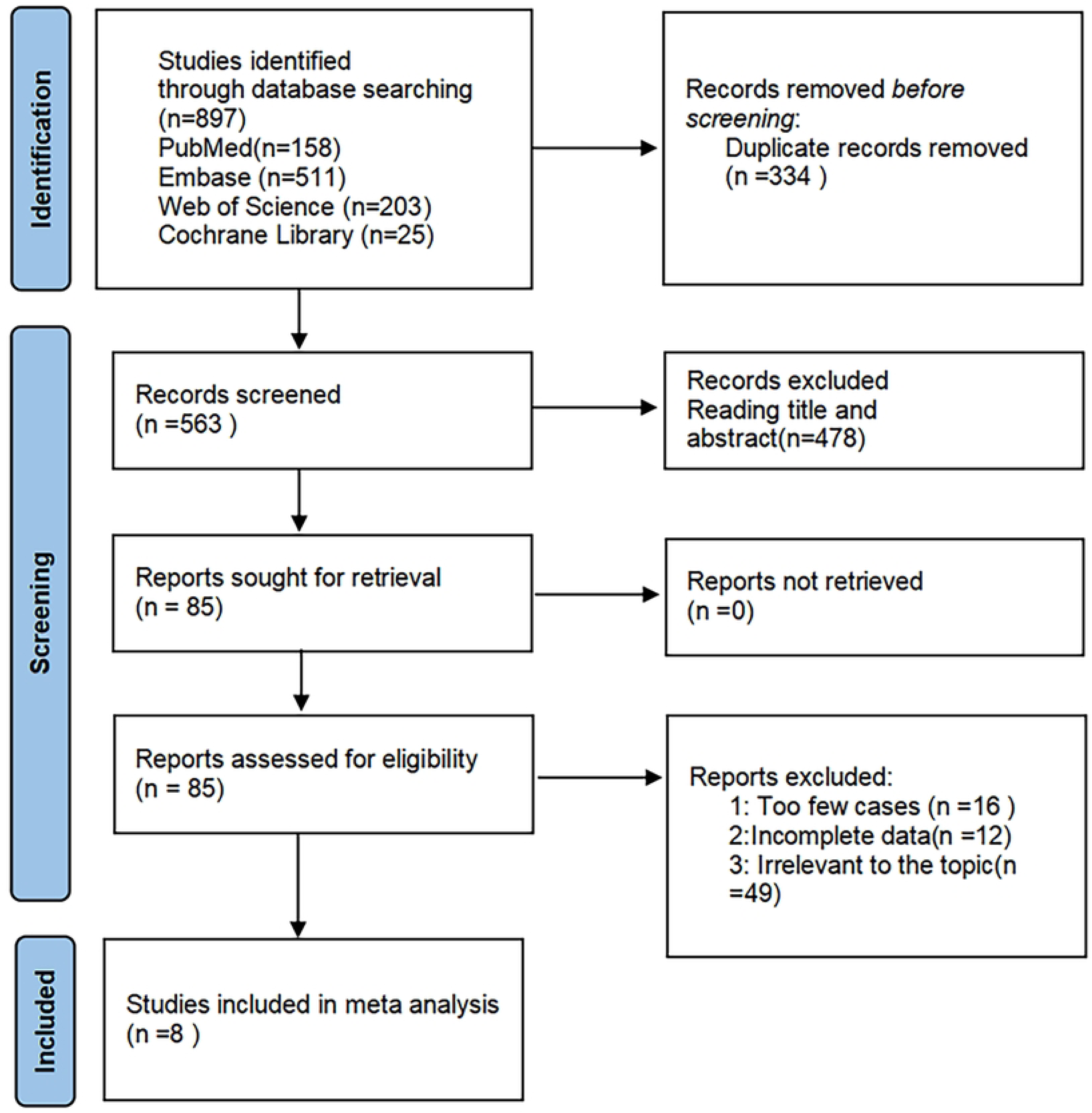

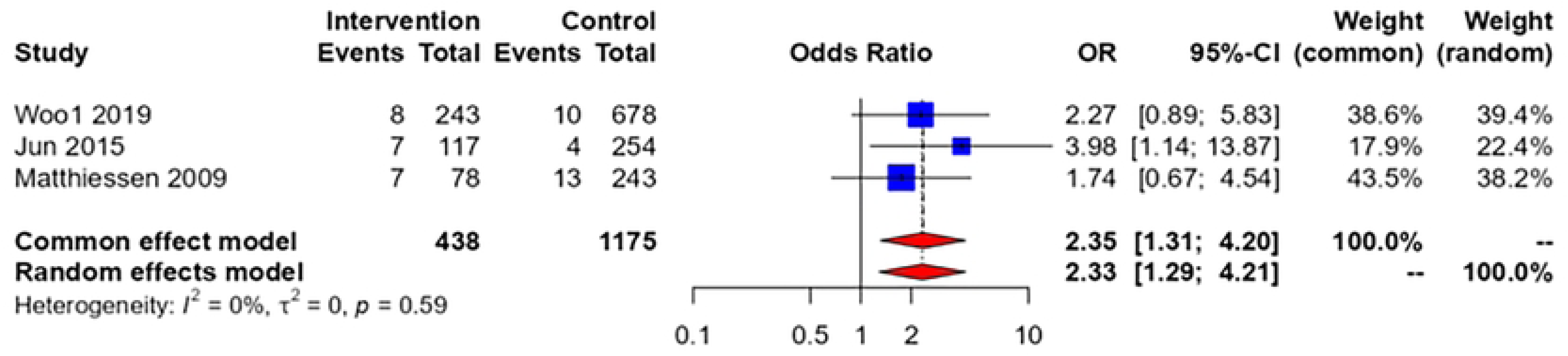

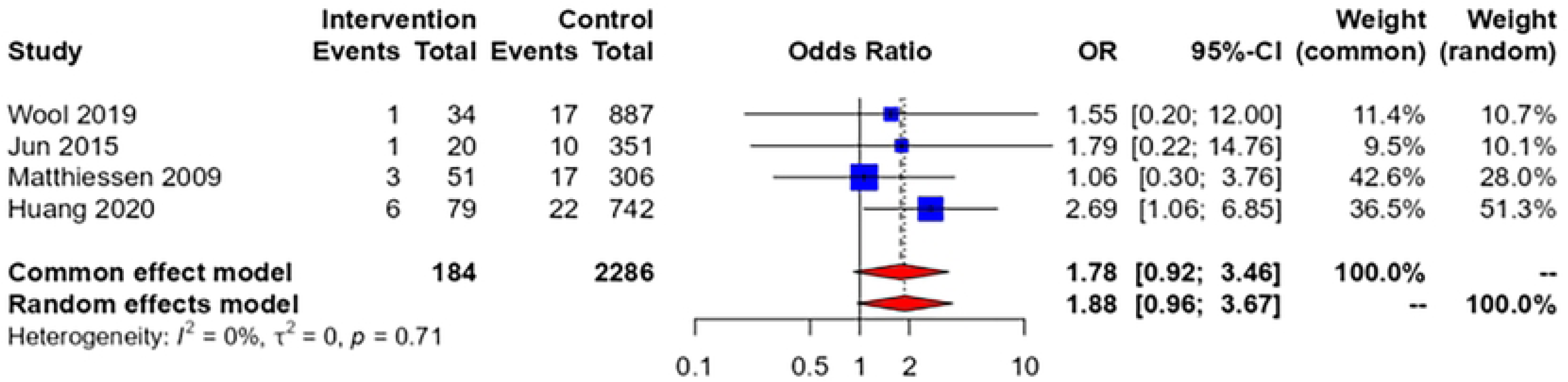

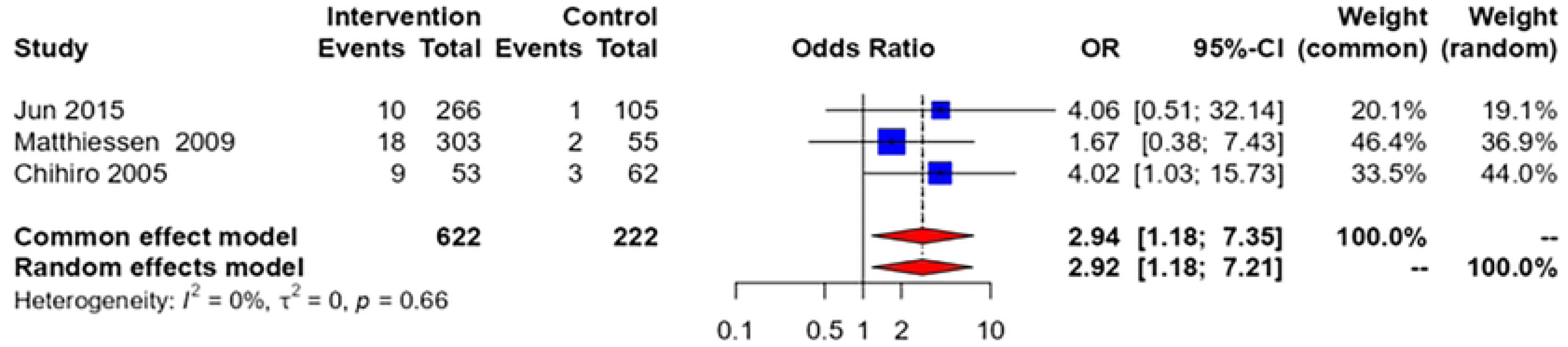

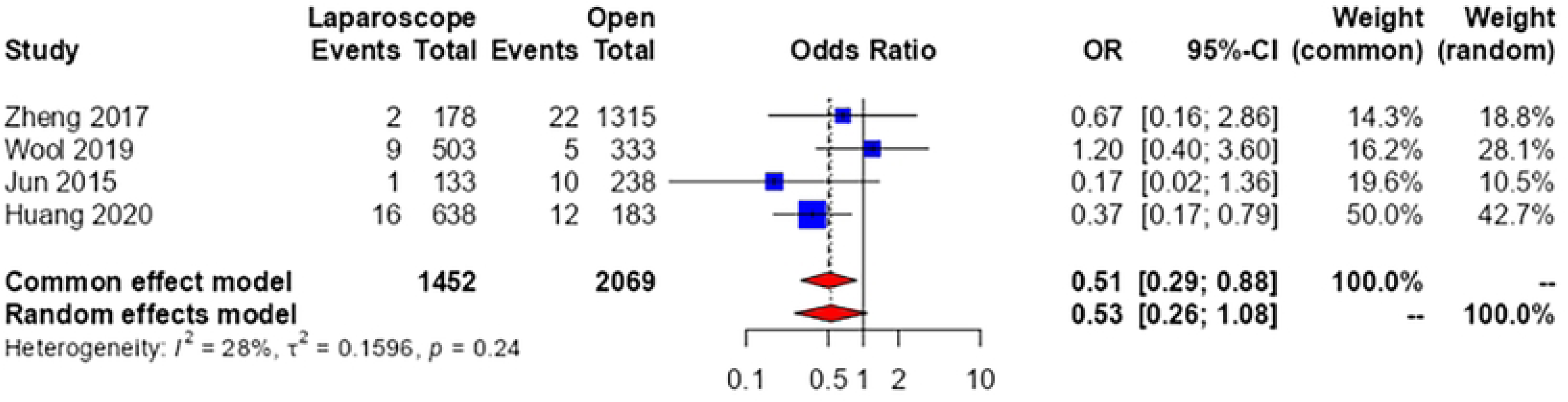

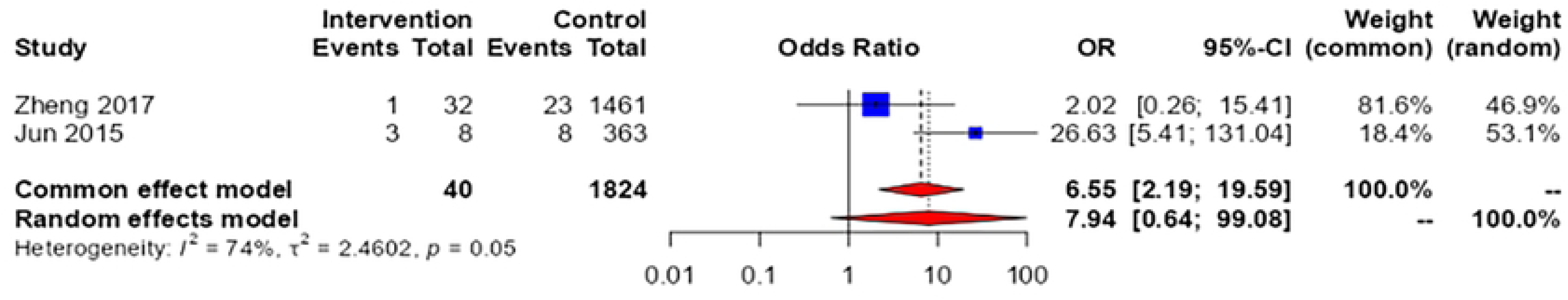

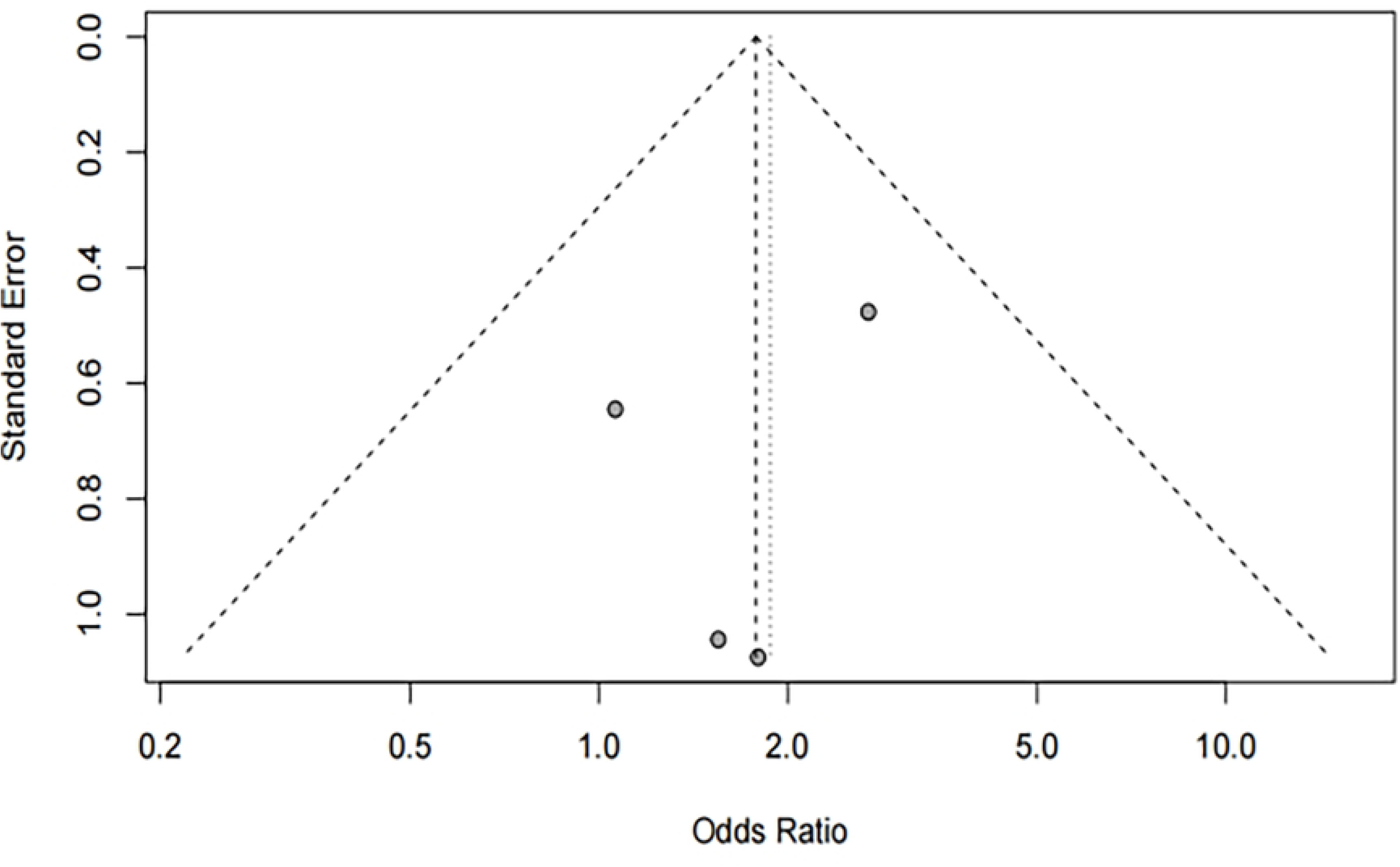

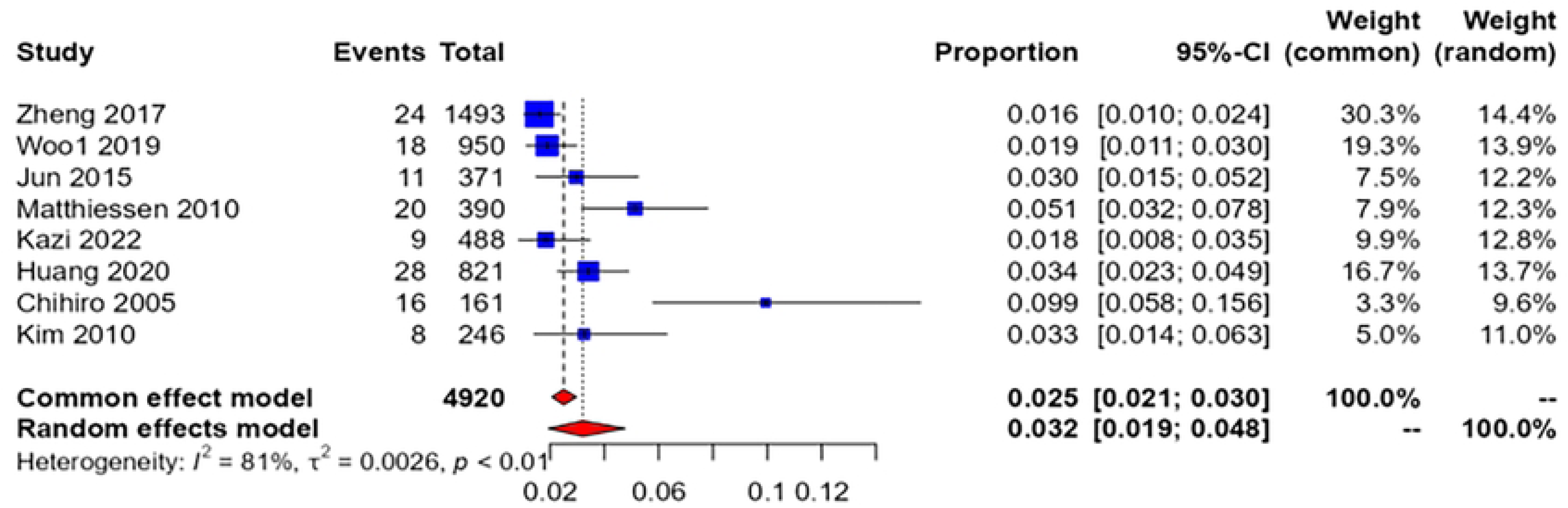

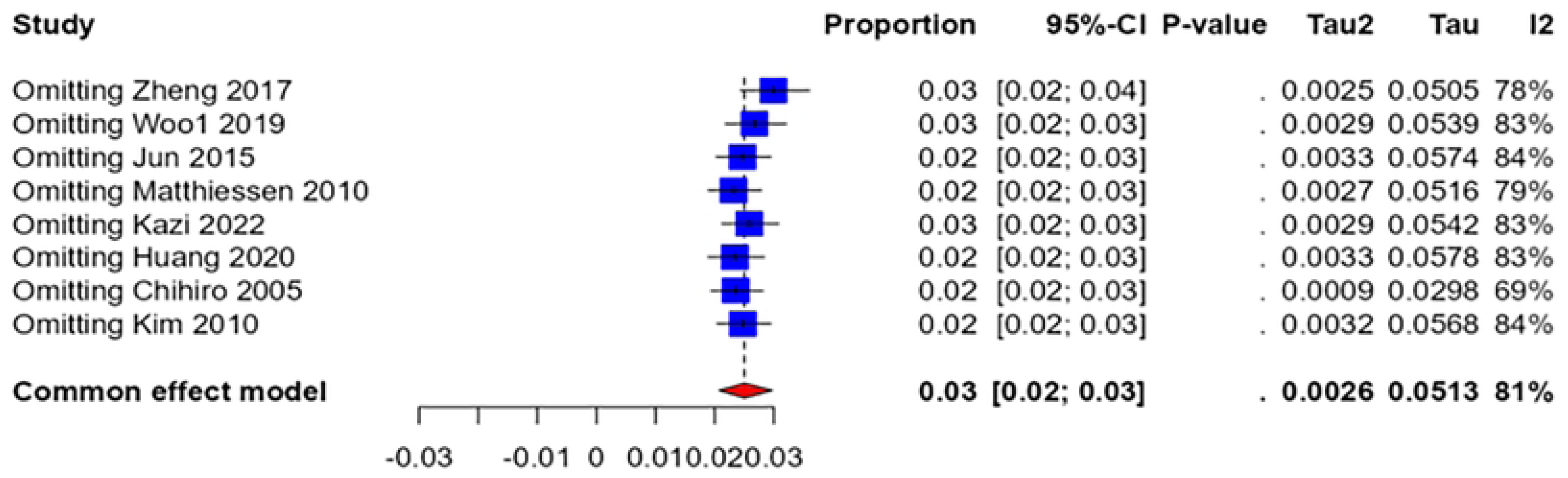

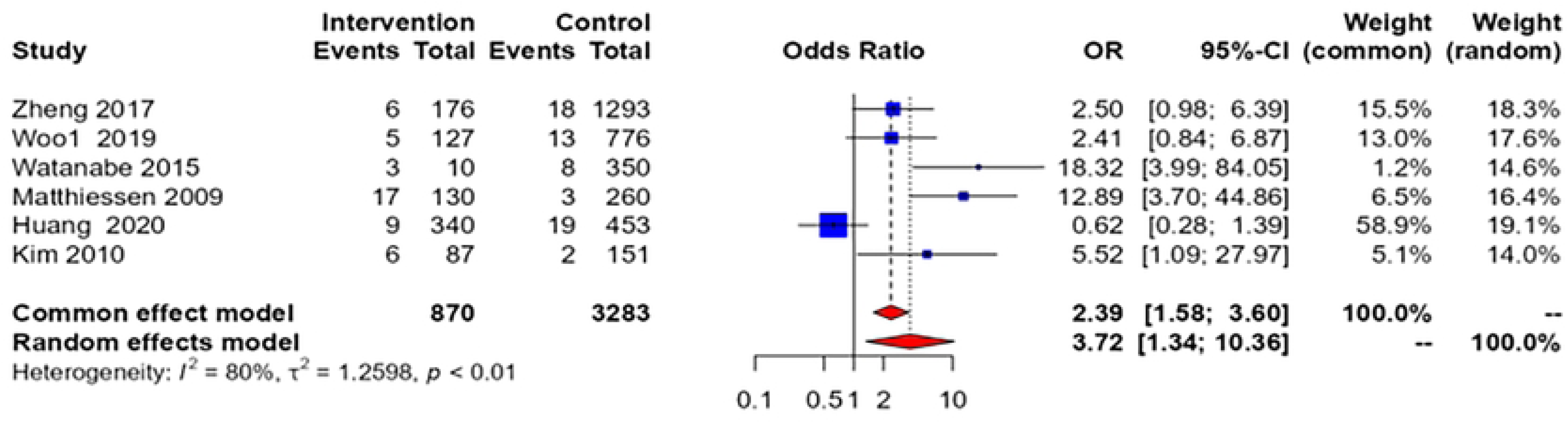

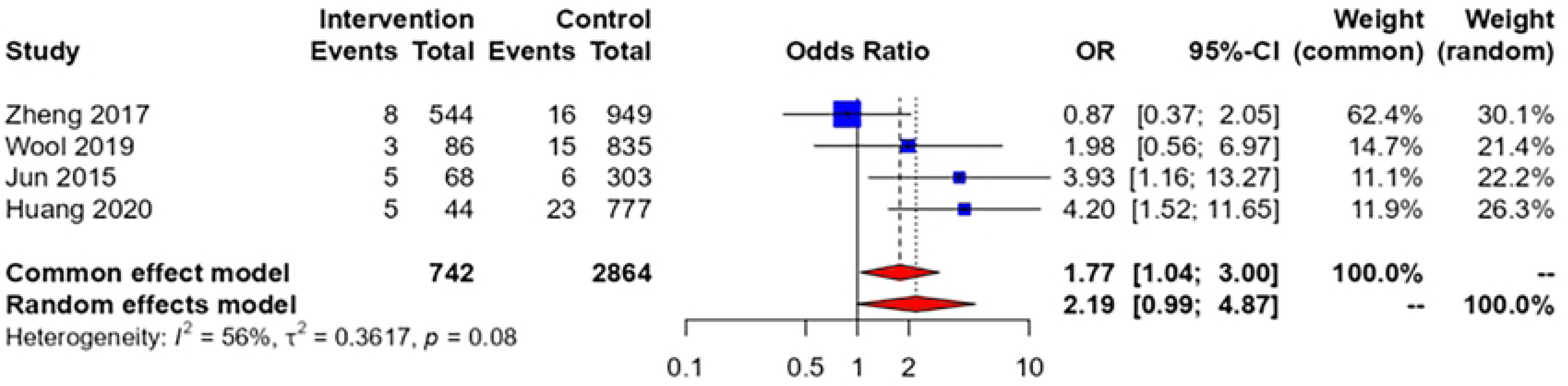

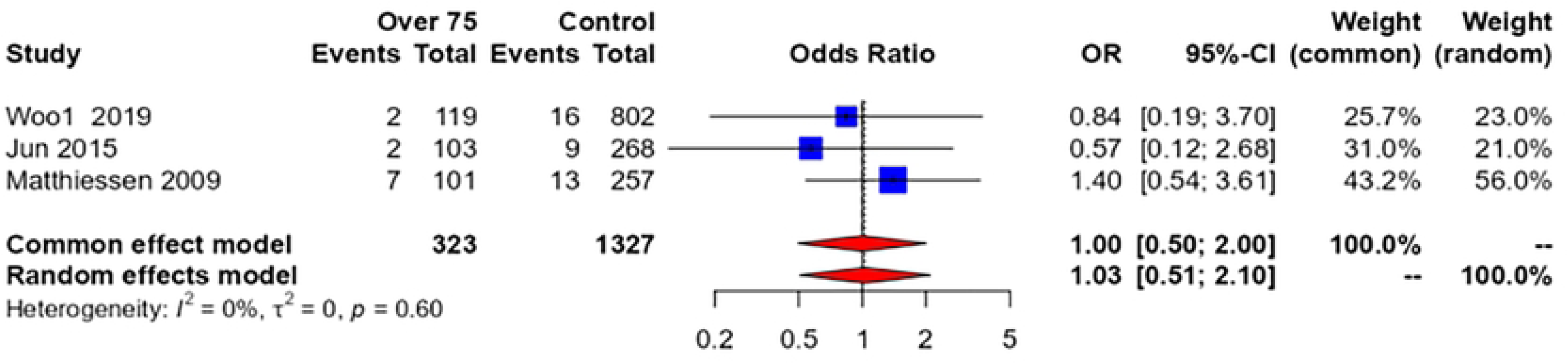

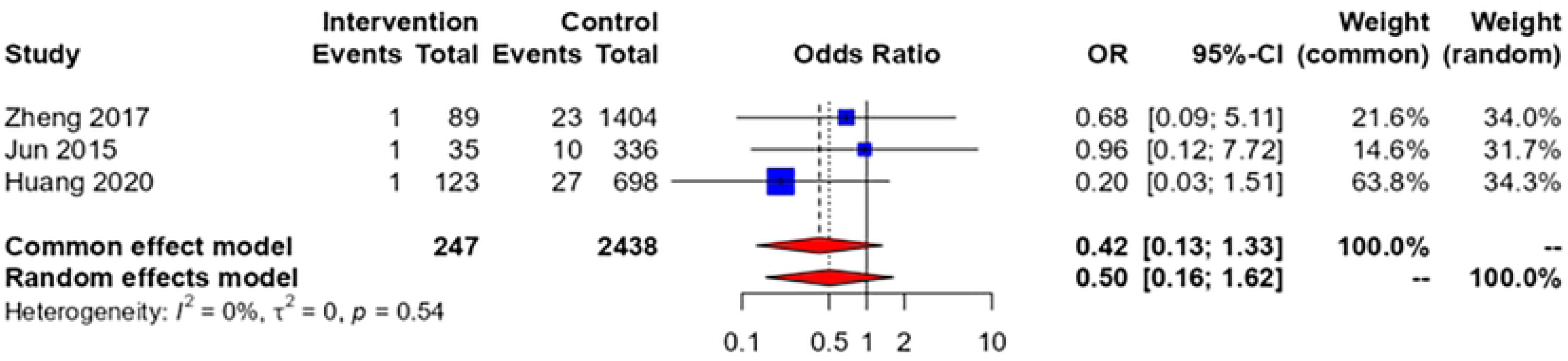

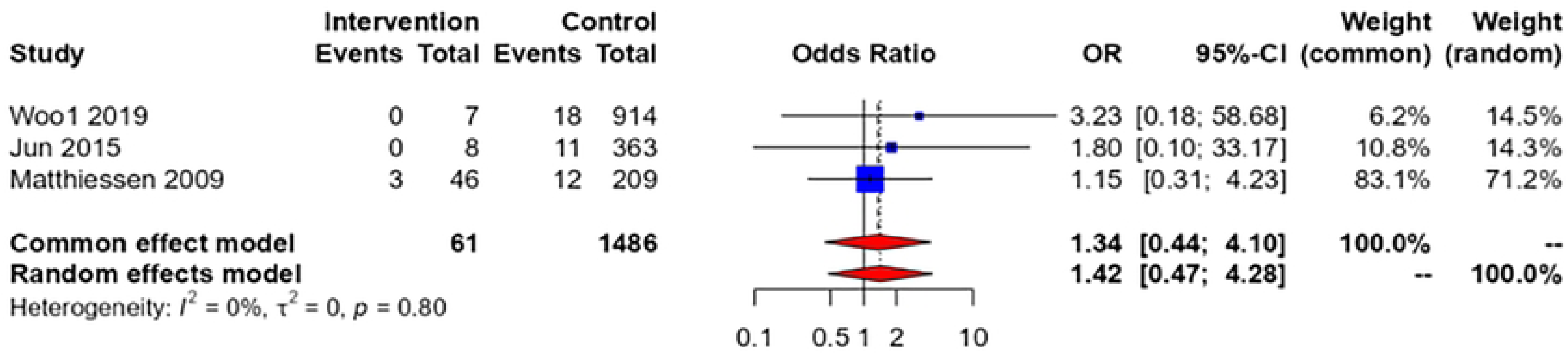

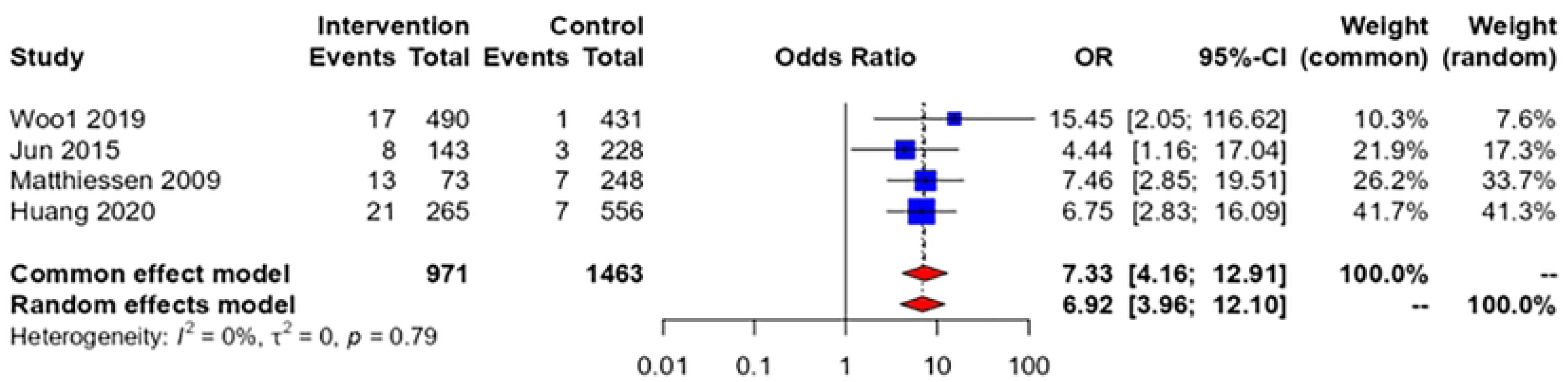

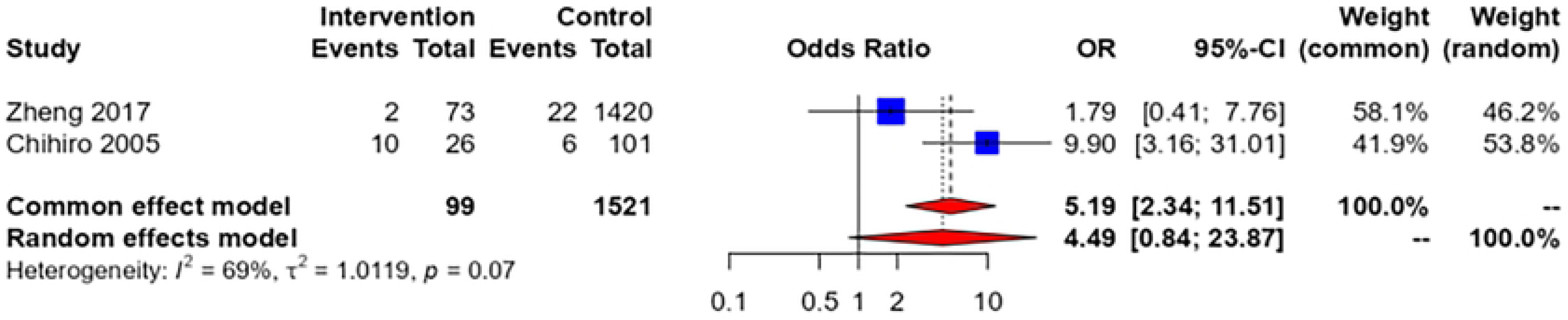

## Notes

### Competing Interest Statement

The authors have declared no competing interest.

### Funding Statement

Yes

### Author Declarations

This study is a meta-analysis. All data are from published literature. The original data are public and do not require ethical or other institutional approval.

